# Associations Between Menstrual Cycle Phases and Heart Rate, Well-Being, and Perceived Exertion in Female Football Players

**DOI:** 10.1101/2024.12.05.24318571

**Authors:** Lucie Lipková, Michaela Beníčková, Adam Wagner, Michal Kumstát

## Abstract

This study aimed to investigate potential associations between menstrual cycle (MC) phases and heart rate (HR), perceived exertion (RPE), and subjective well-being in female football players. Data were collected from 11 players (aged 17–29 years) across 10 competitive matches. HR metrics, subjective well-being, and RPE were monitored in the pre-match, during-match, and post-match periods, as was the menstrual cycle in detail. Key findings revealed that during the early luteal (EL) phase, players spent significantly more time in moderate-intensity HR zones (Zone 4) in both the first (p = 0.046) and second halves (p = 0.051), while spending less time in high-intensity zones (Zone 5) during the first half (p = 0.012). Subjective measures highlighted elevated energy levels in the EL phase compared to the late luteal phase (p = 0.001), underscoring the influence of different menstrual cycle phases on well-being. However, RPE and maximum HR were predominantly shaped by external factors such as opponent strength (p < 0.001) and match outcome (p = 0.035). These results underscore the importance of exploring individual and contextual factors in understanding performance and well-being in female football players.

## Introduction

The participation of women in sports has grown significantly, as demonstrated by the recent Olympic Games, which for the first time featured equal representation of male and female athletes [1]. This trend is particularly evident in women’s football, where the number of officially ranked teams increased from 142 in 2020 to a record 192 by December 2023, highlighting the expanding global influence of women in the sport [2].

Dr. Georgie Bruinvels, a FIFA expert on female sports performance, has emphasized the untapped potential of female athletes: “We still do not know how good females can be. Understanding how to better work with female athletes may unlock new potential, making it an exciting time for women’s football” [3]. This statement highlights the importance of understanding the unique physiological factors that influence female athletes, particularly those that may affect performance on the field.

Performance variability in female soccer players can be influenced by numerous factors, including environmental conditions, surface, quality of the opposition, level of competition, team tactics, nutrition, and sleep [4,5]. Among the most frequently discussed factors influencing performance in female athletes is the menstrual cycle (MC), which involves fluctuations in important hormones such as oestrogen, progesterone, luteinizing hormone, and follicle-stimulating hormone [6]. The MC, typically lasting 21–35 days, is divided into the follicular and luteal phases, separated by ovulation. During the follicular phase, oestrogen levels rise as ovarian follicles mature, peaking before ovulation. In the luteal phase, both oestrogen and progesterone levels increase, with a secondary oestrogen peak in the mid-luteal phase, followed by a decline if fertilization does not occur, leading to menstruation [7,8].

These hormonal changes interact with various physiological systems, such as the cardiovascular, respiratory, metabolic, neuromuscular, and immune functions, and can also affect perceptual responses such as sleep quality, fatigue, and mood [9]. Additionally, thermoregulatory and metabolic processes are influenced, affecting exercise performance through mechanisms like fluid retention, changes in body temperature, and alterations in energy metabolism [10,11]. As a result, the MC can influence both sports performance and overall well-being in female football players [12], but evidence on the MC’s impact on sports performance, particularly in women’s football, remains limited and inconsistent [5,13]. Given these mixed findings, it is recommended that athletes and practitioners monitor MCs and symptoms to tailor training and competition strategies, ultimately improving performance and well-being [5]. Understanding the dynamics of the MC is crucial for enhancing both performance and well-being in female football players.

This study aims to investigate potential associations between of MC phases and heart rate, perceived exertion, and subjective well-being in female football players. By examining these variables, the research seeks to provide insights that could inform the development of training programs and competition strategies tailored to the specific needs of female athletes, ultimately enhancing their performance and health.

## Methods

### Participants

The study initially involved 12 female football players from the Czech Women’s Football League. One player was excluded based on the inclusion criteria (age >15, absence of injuries, non-use of oral contraception, regular MC). The final sample consisted of 11 players (aged 17–29 years), including 2 attackers, 5 midfielders, and 4 defenders. Data collection took place from April to June 2024, encompassing 10 official matches. Measurements were taken before, during, and after each match. All participants provided written informed consent, and for participants under the age of 18, consent was also obtained from their parents or legal guardians. The study protocol was approved by the Ethical Committee (EKV-2024-001).

### Design

This study examined the effects of different MC phases on heart rate, perceived exertion, and subjective well-being in female football players. The research protocol included three assessment stages: pre-match, during the match, and post-match.

- Pre-match: Participants completed a subjective wellness questionnaire 1–2 hours before each match.
- During**-**match: Players’ heart rate (HR) was monitored using wearable sensors throughout the match.
- Post-match: Participants reported their rate of perceived exertion (RPE) and and evaluated the strength of the opponent immediately after the match. Additionally, match outcome and location were recorded to capture relevant contextual factors.

All questionnaires were administered digitally via Microsoft Forms on participants’ mobile devices. At the end of the season, an additional comprehensive questionnaire was used to collect detailed information about participants’ MCs.

### Pre-match Assessment

#### Subjective Wellness Questionnaire

Participants completed a subjective wellness questionnaire (S1 Table) prior to each match to assess their overall well-being based on the previous day’s exertion. The questionnaire covered six factors (energy levels, sleep quality, muscle soreness, diet quality, and stress levels), adapted from Mcgahan et al. [14]. Each item was rated on a five-point Likert scale, where 1 indicated poor well-being and 5 indicated excellent well-being.

### During-match Assessment

#### Heart Rate Monitoring

HR data were collected using the Polar H10 heart rate monitor, integrated with the Polar Team app. The Polar H10 is widely validated and recognized as a gold standard for R-R interval assessments [15,16]. Measurements were recorded throughout the match, including both halves. The key cardiovascular metrics captured were maximal heart rate (HR_max_), mean heart rate, and time spent in the following heart rate zones:

- Zone 1 (Very light): 50–60% of HR_max_
- Zone 2 (Light): 60–70% of HR_max_
- Zone 3 (Moderate): 70–80% of HR_max_
- Zone 4 (Hard): 80–90% of HR_max_
- Zone 5 (Maximum): 90–100% of HR_max_ [17].

HR_max_ was determined as the highest value reached across all recorded data during the matches [18]. This approach was chosen because it reflects the players’ peak physiological response under real match conditions, which is more relevant for assessing in-game performance than values obtained in a controlled test environment.

### Post-match Assessment

#### Rate of Perceived Exertion (RPE)

After each match, participants rated their perceived exertion using a modified category-ratio scale (CR-10 scale), which combines categorical labels with a numerical range (1–10) to quantify perceived exertion. This scale has been validated and shown to be reliable in monitoring training load across various sports [19,20]. The scale (S2 Table) included descriptive labels and smiley faces, with ratings ranging from 1 (lightest possible exertion) to 10 (most strenuous exertion). Ratings were collected 30–60 minutes after the match.

#### Opponent Strength, Match Outcome, and Location

Participants were asked to subjectively evaluate the strength of the opponent after the match, classifying them as weak, medium, or strong, with responses recorded 30–60 minutes after the match via questionnaire. Additionally, match outcome (win, loss, or draw) and match location (home or away) were documented to account for potential contextual factors influencing the players’ performance and perceived exertion.

#### Comprehensive Menstrual Cycle Questionnaire

At the end of the season, participants completed a comprehensive questionnaire (S1 Appendix), which included questions about cycle regularity, the duration of the bleeding phase, and the use of oral contraception. Participants also provided information based on their personal calendars, indicating the days of their bleeding phase from March to June. This data was used to accurately calculate the MC phases and ensure eligibility for the study.

## Menstrual Cycle Phase Calculation

The specific MC phases were identified by tracking the onset of menstrual bleeding over the course of 3 to 4 consecutive cycles for each participant (Table 1).

**Table 1:**
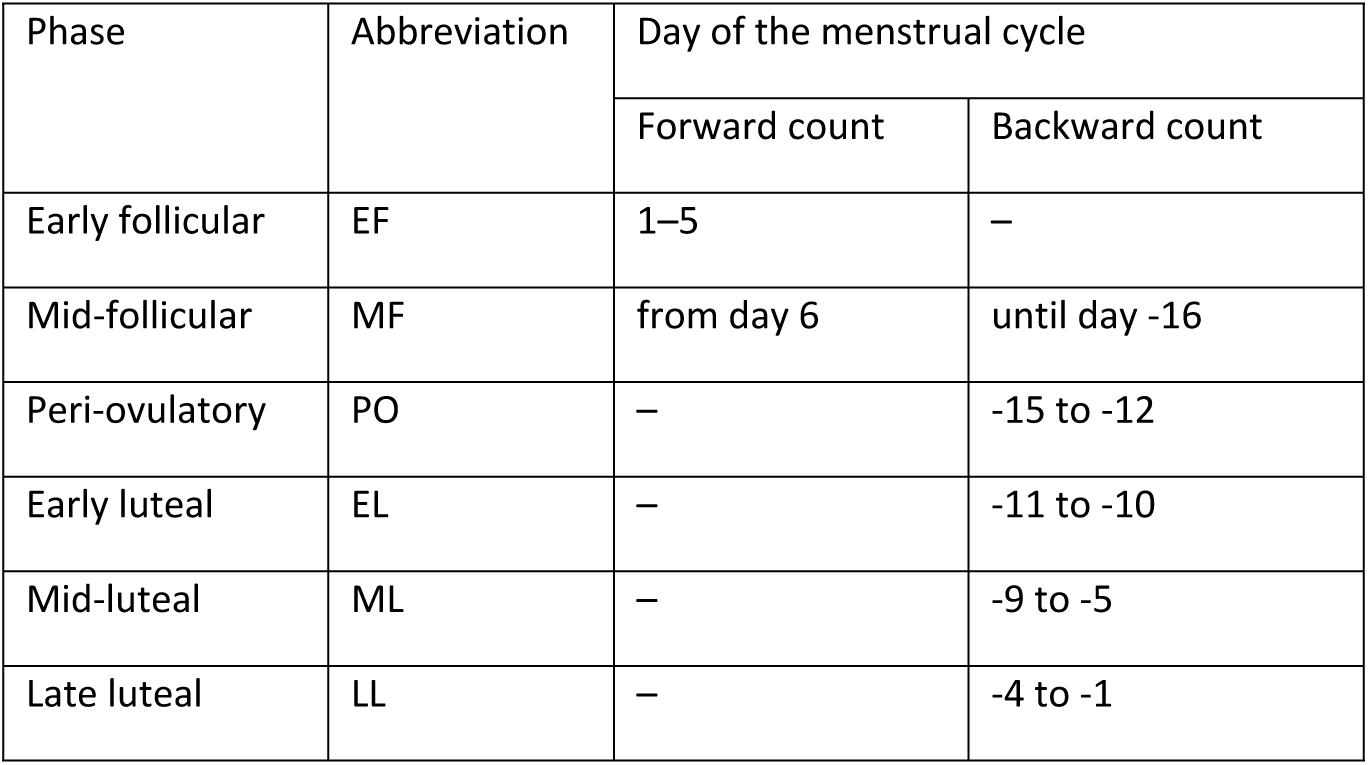
Classification of Menstrual Cycle Phases Based on Forward and Backward Day Count.

The Early Follicular (EF) phase was indicated by the onset of menstrual bleeding and lasted until day 5 of the MC, as described by Elliott-Sale et al. [7]. The Peri-ovulatory (PO) and Mid-luteal (ML) phases were calculated using the counting method outlined by Schmalenberger et al. [8]. Both phases were identified by backward-counting from the first day of the subsequent menstrual bleeding, with the PO corresponding to days −15 to −12, and the ML corresponding to days −9 to −5. The Mid-follicular (MF), Early Luteal (EL), and Late Luteal (LL) phases were assigned to the remaining days of the MC. The MF phase follows the EF phase, beginning on day 6 of the MC and lasting until day −16 (inclusive). The EL phase occurred between the PO and ML phases (days −11 to −10), while the LL phase fell between the ML and EF phases (days −4 to −1).

## Statistical Analysis

Data were analysed using Python within the PyCharm CE IDE. Descriptive statistics were calculated for all variables. Normality was assessed by examining residual histograms, and homogeneity of variance was evaluated by plotting residuals against predicted values. Linear mixed-effects models were employed to evaluate the effects of MC phases and match-related variables (e.g., match location, opposition strength, match result) on dependent measures such as heart rate and subjective wellness scores. Models were estimated using maximum likelihood (ML). The first model assessed heart rate data (maximal heart rate, mean heart rate, and time spent in heart rate zones 1– 5), while the second model focused on subjective metrics (mood, sleep, energy, muscle soreness, diet, stress, and RPE). Both models included fixed effects for MC phase, match result, match location, and opposition quality, with random effects for participant ID to account for repeated measures. Post-hoc comparisons were performed with Holm-Bonferroni corrections to identify significant differences between phases. Pairwise comparisons of residuals from the mixed models were conducted across different MC phases. Effect sizes were estimated using omega squared (ω²) and were interpreted as small (ω² = 0.01), medium (ω² = 0.06), and large (ω² = 0.14) [21].

## Results

### Basic Demographics

The basic demographic characteristics of the participants are summarized in Table 2. All players reported having a regular MC, with an average cycle length of 28.5 days and a bleeding duration of 5.5 days.

**Table 2:**
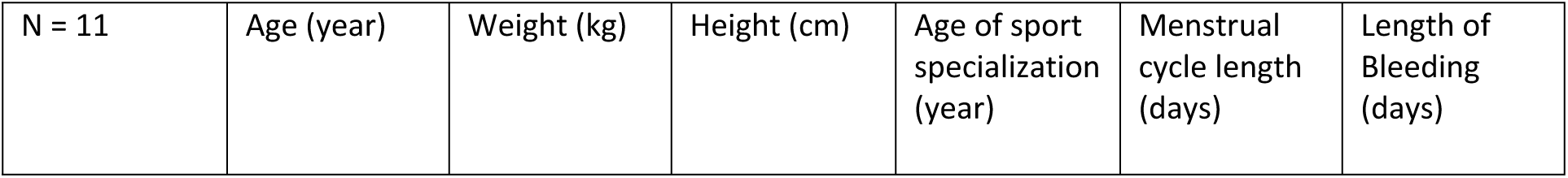
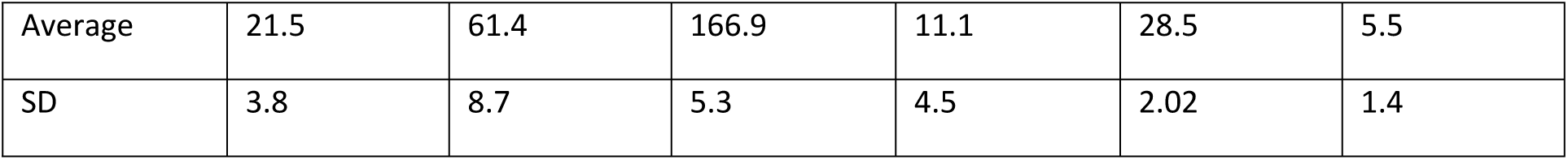
Descriptive characteristics of the players.

### Heart Rate Responses Across Menstrual Cycle Phases

The results for HR variables are displayed in Table 3, with comparisons based on match location, result, and opponent strength, and significant differences detailed in Table 4. Time spent in HR Zone 4 was significantly higher in the EL phase during both the first (p = 0.046, ω² = 0.158) and second halves (p = 0.051, ω² = 0.139). In contrast, time spent in HR Zone 5 in the first half was lower in the EL phase (p = 0.012, ω² = 0.091). Post-hoc tests revealed that the EL phase significantly differed from the LL (p = 0.0491), MF (p = 0.0134), and PO phases (p = 0.019) in time spent in HR Zone 4 – 1st half. Time spent in HR Zone 1 showed significant differences based on opponent strength. Players spent less time in HR Zone 1 during both halves when facing stronger opponents (1st half: p = 0.006; 2nd half: p = 0.037), indicating a higher overall workload in these matches. No other significant differences were identified for the remaining HR parameters across MC phases.

**Table 3:**
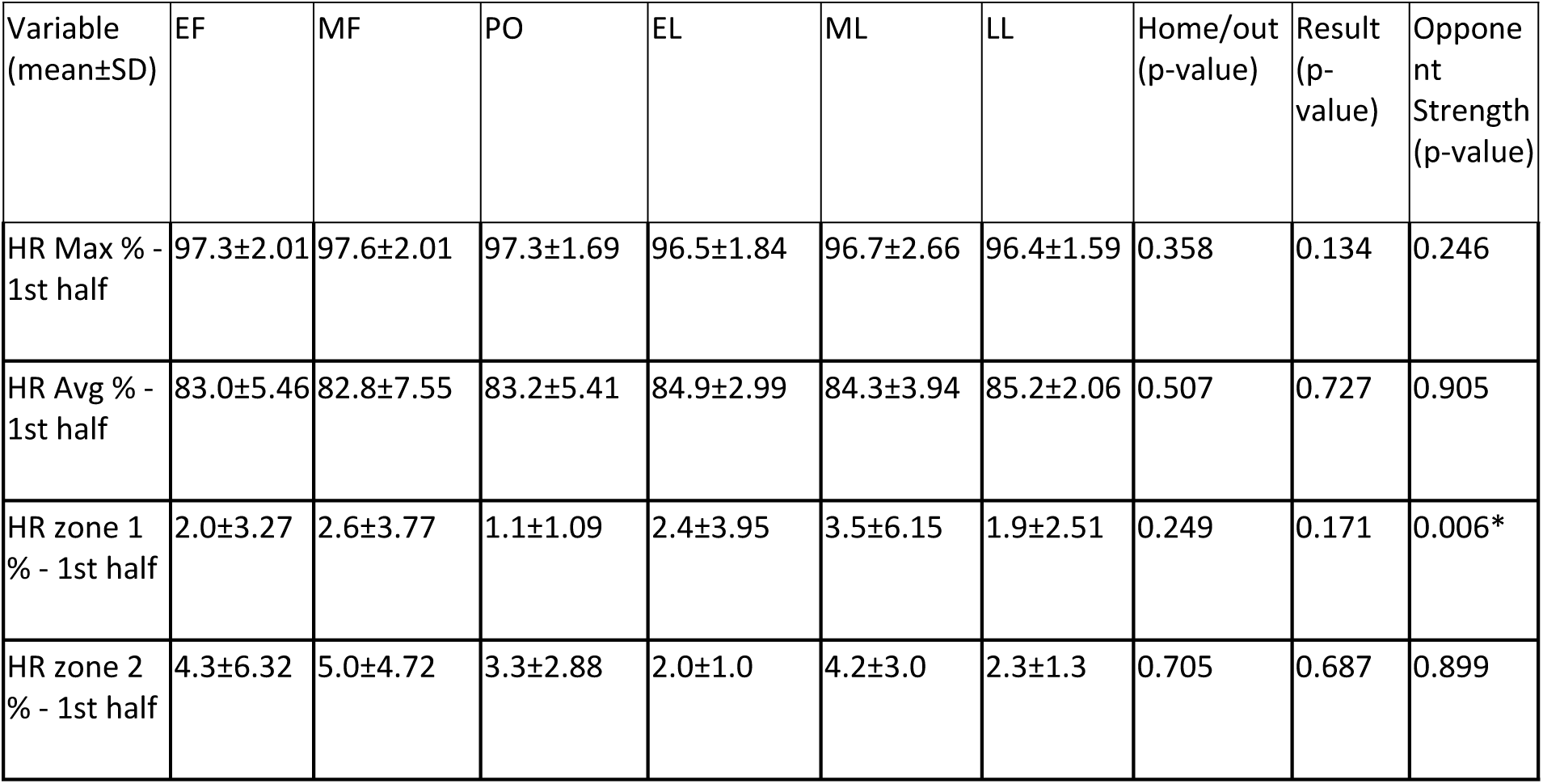
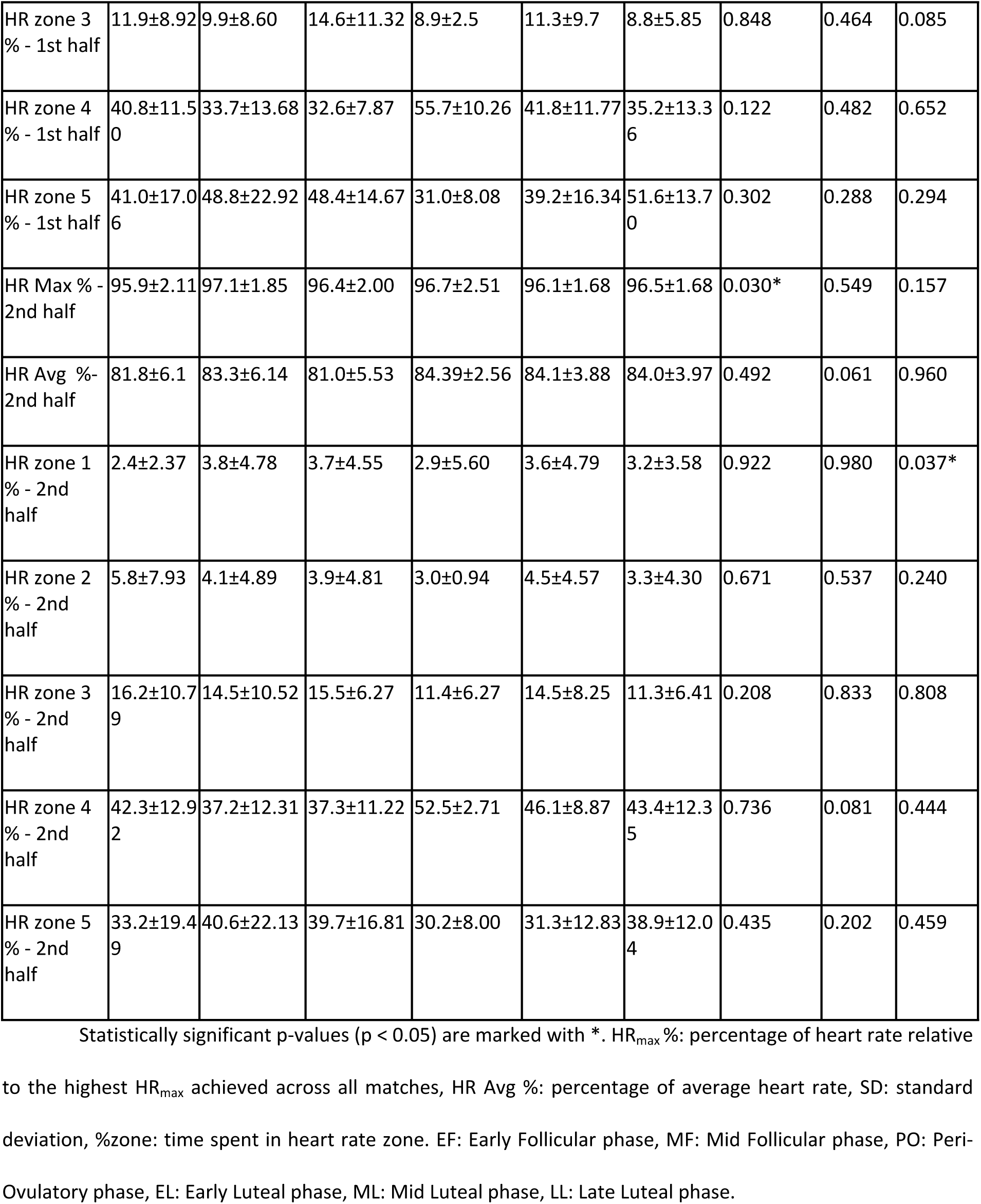
Heart Rate Metrics During the First and Second Halves Across Different MC Phases.

| Variable (mean±SD) | EF | MF | PO | EL | ML | LL | Home/out (p-value) | Result (p-value) | Opponent Strength (p-value) |
| --- | --- | --- | --- | --- | --- | --- | --- | --- | --- |
| HR Max % - 1st half | 97.3±2.01 | 97.6±2.01 | 97.3±1.69 | 96.5±1.84 | 96.7±2.66 | 96.4±1.59 | 0.358 | 0.134 | 0.246 |
| HR Avg % - 1st half | 83.0±5.46 | 82.8±7.55 | 83.2±5.41 | 84.9±2.99 | 84.3±3.94 | 85.2±2.06 | 0.507 | 0.727 | 0.905 |
| HR zone 1 % - 1st half | 2.0±3.27 | 2.6±3.77 | 1.1±1.09 | 2.4±3.95 | 3.5±6.15 | 1.9±2.51 | 0.249 | 0.171 | 0.006* |
| HR zone 2 % - 1st half | 4.3±6.32 | 5.0±4.72 | 3.3±2.88 | 2.0±1.0 | 4.2±3.0 | 2.3±1.3 | 0.705 | 0.687 | 0.899 |
| HR zone 3<br>% - 1st half | 11.9±8.92 | 9.9±8.60 | 14.6±11.32 | 8.9±2.5 | 11.3±9.7 | 8.8±5.85 | 0.848 | 0.464 | 0.085 |
| HR zone 4<br>% - 1st half | 40.8±11.50 | 33.7±13.68 | 32.6±7.87 | 55.7±10.26 | 41.8±11.77 | 35.2±13.36 | 0.122 | 0.482 | 0.652 |
| HR zone 5<br>% - 1st half | 41.0±17.06 | 48.8±22.92 | 48.4±14.67 | 31.0±8.08 | 39.2±16.34 | 51.6±13.70 | 0.302 | 0.288 | 0.294 |
| HR Max % -<br>2nd half | 95.9±2.11 | 97.1±1.85 | 96.4±2.00 | 96.7±2.51 | 96.1±1.68 | 96.5±1.68 | 0.030* | 0.549 | 0.157 |
| HR Avg % -<br>2nd half | 81.8±6.1 | 83.3±6.14 | 81.0±5.53 | 84.39±2.56 | 84.1±3.88 | 84.0±3.97 | 0.492 | 0.061 | 0.960 |
| HR zone 1<br>% - 2nd<br>half | 2.4±2.37 | 3.8±4.78 | 3.7±4.55 | 2.9±5.60 | 3.6±4.79 | 3.2±3.58 | 0.922 | 0.980 | 0.037* |
| HR zone 2<br>% - 2nd<br>half | 5.8±7.93 | 4.1±4.89 | 3.9±4.81 | 3.0±0.94 | 4.5±4.57 | 3.3±4.30 | 0.671 | 0.537 | 0.240 |
| HR zone 3<br>% - 2nd<br>half | 16.2±10.79 | 14.5±10.52 | 15.5±6.27 | 11.4±6.27 | 14.5±8.25 | 11.3±6.41 | 0.208 | 0.833 | 0.808 |
| HR zone 4<br>% - 2nd<br>half | 42.3±12.92 | 37.2±12.31 | 37.3±11.22 | 52.5±2.71 | 46.1±8.87 | 43.4±12.35 | 0.736 | 0.081 | 0.444 |
| HR zone 5<br>% - 2nd<br>half | 33.2±19.49 | 40.6±22.13 | 39.7±16.81 | 30.2±8.00 | 31.3±12.83 | 38.9±12.04 | 0.435 | 0.202 | 0.459 |

Statistically significant p-values ( $p < 0.05$ ) are marked with \*. HR<sub>max</sub> %: percentage of heart rate relative
to the highest HR<sub>max</sub> achieved across all matches, HR Avg %: percentage of average heart rate, SD: standard deviation, %zone: time spent in heart rate zone. EF: Early Follicular phase, MF: Mid Follicular phase, PO: Peri-Ovulatory phase, EL: Early Luteal phase, ML: Mid Luteal phase, LL: Late Luteal phase.

**Table 4:** Statistical Analysis of HR Zone Differences in Female Football Players.

| HR Zone | Comparison of MC Phases | p-value | Omega Squared |
| --- | --- | --- | --- |
| HR Zone 4 - 1st half | EL vs EF | 0.036* | 0.158* |
|  | EL vs MF | 0.001* | 0.158* |
|  | EL vs PO | 0.010* | 0.158* |
|  | EL vs ML | 0.046* | 0.158* |
|  | EL vs LL | 0.023* | 0.158* |
| HR Zone 5 - 1st half | EL vs MF | 0.012* | 0.091 |
|  | EL vs LL | 0.056 | 0.091 |
| HR Zone 4 - 2nd half | EL vs MF | 0.056 | 0.139 |
|  | LL vs MF | 0.051 | 0.139 |
| HR Max % - 2nd half | Home vs Away | 0.030* | 0.166* |
| HR Zone 1 - 1st half | Strong vs Weak Opponent | 0.006* | 0.086 |
| HR Zone 1 - 2nd half | Strong vs Weak Opponent | 0.037* | 0.059 |

Statistically significant p-values ( $p < 0.05$ ) are marked with \*. Strong effect sizes ( $\omega^2 > 0.14$ ) are also marked with \*. EF: Early Follicular phase, MF: Mid Follicular phase, PO: Peri-Ovulatory phase, EL: Early Luteal phase, ML: Mid Luteal phase, LL: Late Luteal phase.

### Subjective Responses Across Menstrual Cycle Phases

The results for subjective response variables are displayed in Table 5, with significant differences detailed in Table 6. Significant differences were found in sleep (p = 0.041, ω² = 0.106), energy (p = 0.003, ω² = 0.136), and stress (p = 0.033, ω² = 0.103. Post-hoc tests revealed that energy levels in the EL phase were significantly higher compared to the LL phase (p = 0.0205). Opponent strength significantly impacted RPE, with players reporting significantly higher perceived exertion against stronger opponents (p < 0.001, ω² = 0.222). No significant differences were found between the different MC phases for RPE.

**Table 5:** Subjective Response Variables Across Different MC Phases.

| Variable<br>(mean±SD) | EF | MF | PO | EL | ML | LL | Home/ou<br>t<br>(p-value) | Result<br>(p-value) | Opponen<br>t<br>Strength<br>(p-value) |
| --- | --- | --- | --- | --- | --- | --- | --- | --- | --- |
| Mood | 3.39±0.96 | 3.66±0.67 | 3.77±0.44 | 3.75±0.50 | 3.56±0.53 | 3.50±0.86 | 0.355 | 0.369 | 0.338 |
| Sleep | 2.31±0.86 | 2.84±0.789 | 3.00±0.58 | 3.25±0.96 | 2.89±0.78 | 3.00±0.88 | 0.127 | 0.499 | 0.813 |
| Energy | 2.77±0.83 | 2.90±0.61 | 3.08±0.28 | 3.75±0.50 | 3.00±0.71 | 2.57±0.76 | 0.441 | 0.457 | 0.846 |
| Muscle<br>Soreness | 3.69±1.03 | 3.68±0.90 | 3.69±0.96 | 4.25±0.96 | 4.00±0.71 | 3.86±0.66 | 0.909 | 0.463 | 0.542 |
| Diet | 2.77±0.83 | 2.84±0.92 | 3.23±0.83 | 3.25±0.50 | 2.89±1.05 | 2.86±0.95 | 0.679 | 0.509 | 0.493 |
| Stress | 2.54±0.66 | 2.87±0.81 | 2.85±0.80 | 3.75±0.96 | 2.67±0.50 | 2.86±0.86 | 0.361 | 0.775 | 0.164 |
| RPE | 6.00±1.35 | 6.18±1.63 | 5.85±0.90 | 6.00±1.63 | 5.67±2.00 | 6.50±1.61 | 0.599 | 0.035* | 0.000* |

Statistically significant p-values ( $p < 0.05$ ) are marked with. SD: standard deviation, EF: Early Follicular phase, MF: Mid Follicular phase, PO: Peri-Ovulatory phase, EL: Early Luteal phase, ML: Mid Luteal phase, LL: Late Luteal phase. Wellness variables were rated on a scale from 1 (poor) to 5 (excellent), and RPE was rated on a scale from 0 (no exertion) to 10 (maximum exertion).

**Table 6:** Statistical analysis results of selected subjective response variables with significant differences in female football players.

| Variable | Comparison of MC Phases | p-value | Omega Squared |
| --- | --- | --- | --- |
| Sleep | EF vs MF | 0.041* | 0.106 |
|  | EF vs PO | 0.013* | 0.106 |
|  | EF vs LL | 0.028* | 0.106 |
| Energy | EL vs EF | 0.003* | 0.136* |
|  | EL vs MF | 0.004* | 0.136* |
|  | EL vs PO | 0.056 | 0.136* |
|  | EL vs ML | 0.017* | 0.136* |
|  | EL vs LL | 0.001* | 0.136* |
|  | PO vs LL | 0.035* | 0.136* |
| Stress | EL vs EF | 0.033* | 0.103 |
|  | EL vs ML | 0.057 | 0.103 |
| RPE | Home vs Away | 0.599 | 0.222* |
|  | Result (Win vs Lose) | 0.035* | 0.222* |
|  | Strong vs Weak Opponent | < 0.001* | 0.222* |

Statistically significant p-values ( $p < 0.05$ ) are marked with \*. Strong effect sizes (omega squared > 0.14) are also marked with \*. EF: Early Follicular phase, MF: Mid Follicular phase, PO: Peri-Ovulatory phase, EL: Early Luteal phase, ML: Mid Luteal phase, LL: Late Luteal phase.

## Discussion

This study investigated associations between the different phases of the MC and HR during matches, as well as subjective well-being in female football players. A key finding (Fig 1) was that during the EL phase, players were observed to spend more time in HR Zone 4 (moderate-intensity exercise) and less time in HR Zone 5 (high-intensity exercise), while reporting higher energy levels. This suggests that players may be better suited for moderate-intensity efforts during the EL phase, while their capacity for high-intensity performance may be reduced. In contrast, no significant differences were observed across the other MC phases, suggesting a limited and phase-specific impact of the menstrual cycle on HR and subjective responses.

**Fig 1:**
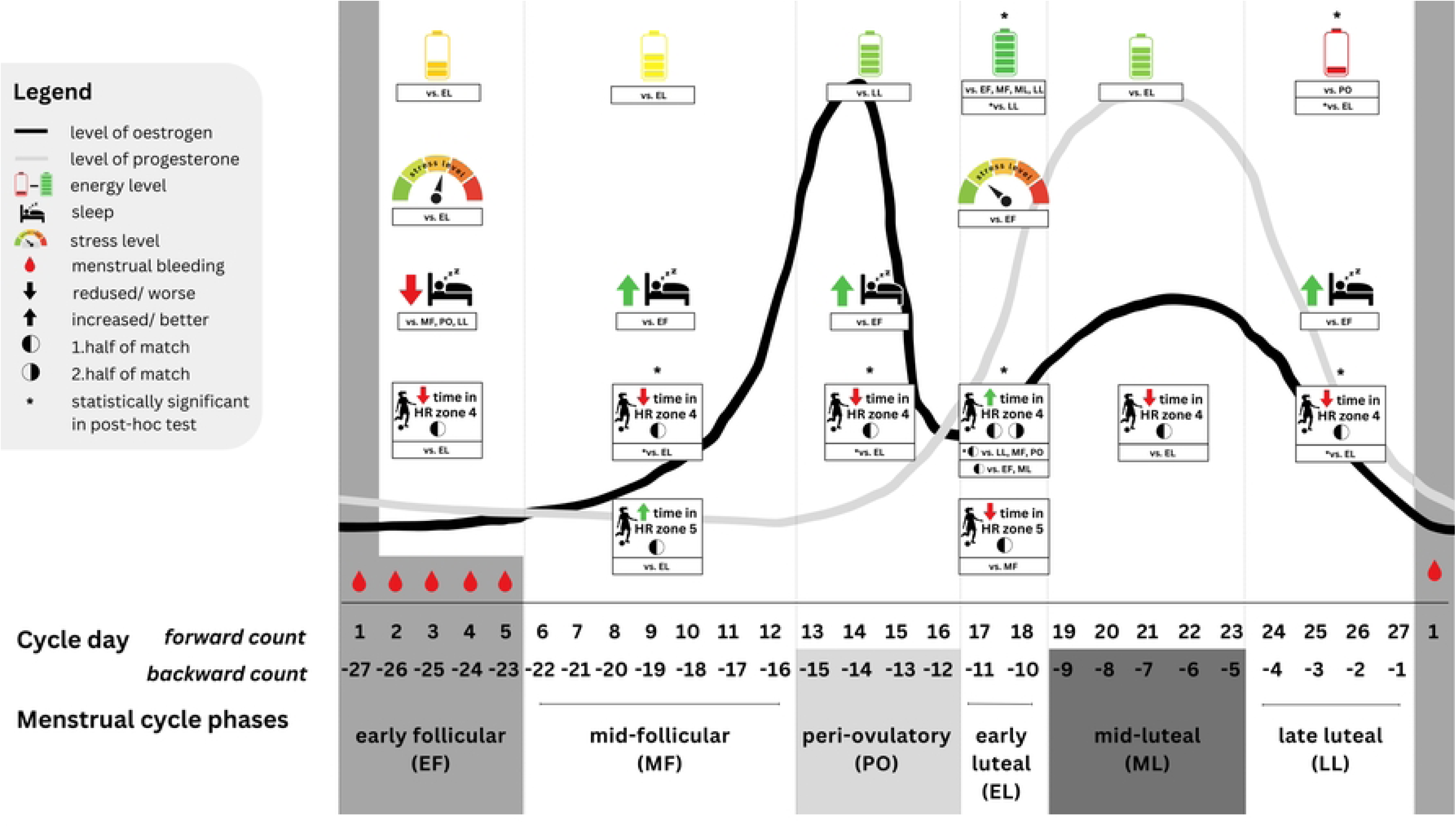
Summary of findings.

### Heart Rate and Menstrual Cycle Phases in Female Football Players

The observed HR responses may be associated with typical hormonal fluctuations, particularly the rise in progesterone and relatively elevated oestrogen levels during the EL phase. Progesterone is known to increase HR by elevating sympathetic nervous system activity, influencing vascular tone and potentially limiting the heart’s ability to reach higher intensity HR zones [22]. Meanwhile, oestrogen’s vasodilatory effects [23,24] improve aerobic performance by reducing peripheral resistance and increasing cardiac output, contributing to enhanced moderate-intensity performance during the EL phase. Based on our findings, differences in HR responses between MC phases may be linked to these hormonal variations, with players showing a preference for moderate-intensity efforts (HR Zone 4) during the EL phase, while time in high-intensity efforts (HR Zone 5) was reduced. Together, these hormonal effects suggest that the balance between progesterone and oestrogen during the luteal phase may optimize moderate-intensity performance while simultaneously challenging the ability to sustain high-intensity efforts. Further investigation into these mechanisms is needed to fully understand how progesterone and oestrogen interact to modulate HR during different phases of the MC [11].

These findings are consistent with previous research, which highlights increased cardiovascular load and HR during the luteal phase [25]. This makes it more difficult for athletes to achieve higher HR zones during submaximal exercise, corroborating our findings of increased time spent in HR Zone 4 during the EL phase. Similarly, enhanced moderate-intensity exercise capacity has been reported across various sports during the luteal phase, including football and futsal [22,26]. The consistency of these findings underscores the significant role of hormonal fluctuations in modulating cardiovascular and performance metrics during the MC.

However, not all studies align with these findings. Some research, such as McNulty et al. [27], found no significant differences in anaerobic capacity or maximal HR across MC phases, suggesting that while HR variability may be sensitive to hormonal changes, it does not always translate into differences in sprint or high-intensity performance. Wiecek (2019) [28] emphasized that cardiovascular responses may vary with the MC, but their impact on performance is influenced by factors like fitness level, training adaptations, and the specific demands of the sport. This variability highlights the complexity of hormonal interactions across MC phases and the need for further research, particularly involving detailed hormonal profiling, to clarify the practical implications of MC phases on female football players’ performance.

While hormonal fluctuations clearly play a significant role in modulating HR responses, our study also highlighted the influence of external factors, such as opponent strength and home advantage. Players spent less time in HR Zone 1 (low intensity) when competing against stronger opponents, indicating a higher overall workload in these matches. Additionally, HR_max_ in the second half of matches was significantly higher when playing at home, which may reflect the psychological and environmental advantages of playing in familiar surroundings. Although differences in HR across MC phases were observed, no significant differences were found for HR_max_ between phases. This suggests that maximum performance may be more influenced by external factors, such as match outcome or home advantage, than by MC phases alone. Overall, our findings highlight that the EL phase, alongside external factors, may play a significant role in modulating cardiovascular responses and performance in female football players.

### Subjective Responses Across Menstrual Cycle Phases

In this study, significant differences in subjective responses were observed across MC phases, particularly in sleep, energy, and stress. Energy levels were higher during the EL phase compared to the LL phase, consistent with research linking hormonal changes, such as elevated oestrogen levels, to enhanced mood and energy [20]. Additionally, RPE was significantly impacted by external factors, with higher exertion reported when playing against stronger opponents and in matches that were lost. However, no significant differences in RPE were detected between the MC phases, suggesting that while certain subjective variables may vary across phases, perceived exertion is more responsive to the competitive context than by MC phases themselves.

#### Energy Levels

Energy levels fluctuated significantly across MC phases, with the lowest values reported during the LL and EF phases. These findings may reflect the influence of typical hormonal changes— particularly variations in oestrogen and progesterone—on serotonin levels, which play a key role in regulating mood and energy. The higher energy levels observed in the EL phase could be associated with elevated oestrogen levels during the late follicular phase, known to enhance serotonin transmission [20,29]. Although post-hoc tests did not confirm significant differences across all phases, the variation in energy levels may be due to individual hormone sensitivity or external factors affecting energy perception, such as the accuracy of menstrual cycle tracking.

#### Sleep

Sleep, another critical factor influencing well-being, also varies across the MC. Previous research shows that females are more prone to sleep disturbances than males, which can negatively affect athletic performance, increase perceived effort, and impair cognitive and motor function [29]. In our study, sleep quality fluctuated across MC phases, with no significant differences found in post-hoc analyses. Some research suggests more sleep disturbances in the LL phase, while others report no significant differences in sleep, fatigue, or soreness across phases [30], highlighting the variability in the influence of the MC on sleep.

#### Rate of Perceived Exertion (RPE)

In our study, no significant differences in RPE were observed across MC phases, consistent with prior research [9]. Similarly, Abbott et al. [19] found no effect of MC phase on session RPE (sRPE)— which refers to the overall perceived exertion for an entire training session—or external load during matches. Although it has been hypothesized that rising progesterone levels during the luteal phase may increase perceived effort, our findings did not support this assumption, aligning with other studies [31,32].

However, some studies do report higher sRPE during the luteal phase, particularly for longer or more intense activities. For instance, sRPE was found to be higher for distances over 5 km during the luteal phase compared to menstruation, and players reported greater fatigue 48 hours post-match in the luteal phase than in the follicular phase [30]. This variation suggests that while MC phases may not significantly influence perceived effort during shorter or less intense activities, they could play a more pronounced role in longer, more demanding exercises.

As expected, RPE in our study was significantly influenced by external factors, with higher perceived exertion reported when playing against stronger opponents and in matches that were lost. These findings underscore the interaction between external variables and perceived effort, highlighting that match conditions and exertion demands may have a more pronounced impact than the MC phases themselves.

#### Stress and Other Well-Being Factors

Although stress levels varied across MC phases—peaking during the EF and ML phases and lowest during the EL phase—post-hoc tests did not confirm significant differences. This suggests that while there may be fluctuations in stress associated with hormonal changes, our findings did not provide strong evidence to support this relationship. Previous research has linked hormonal fluctuations to mood and stress regulation [20]. Other well-being factors, including mood, muscle soreness, and diet, showed no significant phase-related differences.

The luteal phase is often associated with premenstrual syndrome (PMS), characterized by symptoms like sleep disturbances, anxiety, fatigue, and water retention that negatively affect well-being [29]. Increased progesterone during this phase may contribute to mood disturbances and muscle soreness due to alterations in neurotransmitter systems and amino acid metabolism [30]. Our study found that pre-match well-being did not significantly affect match outcomes or perceptions of opponent strength, indicating that subjective feelings before the match had no influence on performance or opponent perception.

### Practical Application

This study emphasizes the complex interplay between MC phases, external factors, and performance in female football players. While associations between the early luteal phase and moderate-intensity efforts were observed, future research should investigate potential causal links. Practical strategies could involve monitoring MC phases to inform individualized training adaptations during the EL phase, particularly for moderate-intensity efforts. Additionally, external situational factors, such as opponent strength and match demands, should guide recovery and load management to support optimal performance.

## Limitations

This study relied on calendar-based tracking to estimate MC phases, without objective hormonal measurements. As a result, we could not distinguish between ovulatory and anovulatory cycles or luteal phase deficiencies, potentially introducing variability into the findings [33]. Given the high activity levels of our participants, it is likely that some experienced anovulatory or luteal phase-deficient cycles despite regular menstruation [11]. In such cases, hormone levels, particularly progesterone, would remain low [7,11]. This highlights the need for hormonal profiling in future research to improve accuracy.

Additionally, football performance is shaped by numerous external factors, such as pitch quality, player position, weather, team tactics, and psychological stress. These variables, along with match-specific elements like opponent strength and injuries, add significant variability to both performance and perceived exertion. These complexities underscore the challenge of assessing physiological responses in live football settings and should be considered when interpreting results.

## Conclusion

This study highlights the combined influence of MC phases and external factors on performance and well-being in female football players. While the EL phase was associated with improved moderate-intensity performance and higher energy levels, external factors, such as opponent strength and match outcomes, had a more pronounced effect on low intensity performance, maximum HR and RPE. These findings emphasize the need to integrate both internal physiological states and situational demands into training and performance strategies. By leveraging an ecologically valid approach, this research provides actionable insights to optimize performance in real-world conditions.

## Data Availability

All relevant data are within the manuscript and its Supporting Information files.

## Supporting Information

**S1 Table. This is the S1 Table Wellness Questionnaire**

**S2 Table. This is the S2 Table RPE scale**

**S1 Appendix. This is the S1 Appendix file.**

**S1 File. Raw data in excel worksheet.**

